# Consistency of automated coronary calcium score and extent of emphysema with different CT scanners and radiation dose protocols in lung cancer screening

**DOI:** 10.1101/2024.01.10.24301102

**Authors:** Roberta Eufrasia Ledda, Gianluca Milanese, Maurizio Balbi, Federica Sabia, Camilla Valsecchi, Margherita Ruggirello, Nicola Sverzellati, Alfonso Vittorio Marchianò, Ugo Pastorino

## Abstract

**Objective:** To assess the consistency of automated measurements of coronary artery calcification (CAC) burden and emphysema extent on computed tomography (CT) images acquired with different scanners and radiation dose protocols in a lung cancer screening (LCS) population.

**Materials and Methods:** The analysis included two LCS cohorts, named *inter-scanner cohort*, whose subjects underwent two consecutive screening rounds with two different dual-source CT scanners, and *inter-dose cohort*, whose subjects underwent a low-dose CT scan and an ultra-low dose CT scan.

Exclusion criteria for CAC measurements were software failure, previous history of CVD and/or of coronary stenting, whereas for emphysema assessment software failure only. CT images were retrospectively analyzed by a fully automated AI software for CAC scoring, using three predefined Agatston score categories (0-99, 100-399, and ≥ 400), and emphysema quantification, using the percentage of low attenuation areas (%LAA). Demographic and clinical data were obtained from the written questionnaire completed by each participant at the first visit.

Agreement for CAC and %LAA categories was measured by the k-Cohen Index with Fleiss-Cohen weights (K_w_) and 95% Confidence Interval (CI).

**Results:** In the inter-scanner cohort, an overlap of CAC strata was observed in 218/245 (90%) volunteers with an almost perfect agreement (K_w_= 0.91, 95%CI 0.88-0.95), while an overlap of %LAA strata in 182/256 (71%) volunteers, with a substantial agreement (K_w_= 0.70, 95%CI 0.63-0.76). In the inter-dose cohort, an overlap of CAC strata was observed in 275/327 (84%) volunteers, with an almost perfect agreement (K_w_= 0.86, 95%CI 0.82-0.90), while an overlap of %LAA strata was found in 204/356 (57%) volunteers, with a moderate agreement (K_w_= 0.57, 95%CI 0.51-0.63).

**Conclusion:** Automated CAC and emphysema quantification showed consistent results when applied on CT images acquired with different scanners and different radiation dose CT protocols in two LCS cohorts.

## Introduction

Lung cancer (LC), chronic obstructive pulmonary disease (COPD), and cardiovascular disease (CVD) – “the Big-3” – are expected to be the leading causes of death by 2050 globally [1]. Besides pulmonary nodules, low dose computed tomography (LDCT) of the chest allows detection and quantification of coronary artery calcification (CAC) and pulmonary emphysema, recognized biomarkers for CVD and COPD, respectively [2, 3].

Driven by the evidence of a 20-40% reduction of LC mortality [4, 5], several national stakeholders and international scientific societies have been increasingly endorsing LDCT-based lung cancer screening (LCS) programs [6-9]. With the Big-3 sharing their main risk factors [1], these efforts toward LCS implementation might offer a unique opportunity to improve CVD and COPD secondary prevention through a large availability of LDCT images. Such a potentially huge number and heterogeneity of imaging data pose the need for a timesaving and reproducible approach for imaging analysis. The radiological literature explored the field of quantitative imaging, leading to the availability of objective parameters for assessing both emphysema and CAC, mostly exploiting Artificial Intelligence (AI) – based systems [10-12]. Nonetheless, whether and to what extent an objective and reproducible AI-based quantification of CAC and emphysema is affected by CT equipment and radiation dose is still to be defined.

This retrospective study aimed at assessing the consistency of automated measurements of CAC score and emphysema extent on CT images of two LCS cohorts, acquired with two different scanners and with two different radiation dose CT protocols, namely, LDCT and ULDCT, using a commercially available AI software.

## Materials and Methods

### Study population and imaging acquisition

This retrospective study was performed on prospective data acquired from the bioMILD trial (clinicaltrials.gov ID: NCT02247453), an ongoing prospective study testing the combination of plasma miRNA and LDCT to improve the efficacy of LCS by individual risk profiling and personalized screening intervals. Details on the bioMILD trial have been described elsewhere [13]. The original Institutional Review Board approval and written informed consent allowed the use of data for future research.

The present analysis included two cohorts extracted from the bioMILD population:

a. A first cohort (hereafter named inter-scanner cohort) comprised 259 screenees who underwent two consecutive screening rounds with two different dual-source CT scanners. For each participants the first scan was performed with a second-generation dual-source CT scanner (Somatom Definition Flash, Siemens Medical Solutions) while the second scan with a third generation dual-source CT scanner (Somatom Force, Siemens Healthineers, Erlangen, Germany). Scanning parameters are detailed in the *Supplementary materials*.
b. A second cohort (hereafter named inter-dose cohort) comprised 361 consecutive screenees on whom an incident screening round was performed between February and July 2019. LDCT scans were obtained with a third generation dual-source CT scanner (Somatom Force, Siemens Healthineers, Erlangen, Germany); each participant underwent double scan in a single breath-hold: one LDCT scan and one ultra-low dose CT (ULDCT). A detailed description has been reported elsewhere [14], while scanning parameters are described in the *Supplementary materials*.

### Clinical data

Data on past medical history were collected through dedicated questionnaires and direct interviews with a study investigator at the baseline screening round.

### CAC and Emphysema quantification

LDCT and ULDCT images were transferred to a dedicated workstation (Alienware Area 51 R6 equipped with Dual NVIDIA GeForce RTX 2080 OC graphics) and analyzed by a fully automated AI software (AVIEW, Coreline Soft). As reported elsewhere [10], CAC was measured with a 3-dimensional U-net architecture-based scoring tool and stratified using three predefined Agatston score categories: 0-99, 100-399, and ≥ 400 [15]. Emphysema was quantified using the percentage of lung volume occupied by voxels with attenuation ≤ −950 Hounsfield Units, HU (percentage of low attenuation areas, %LAA) [16, 17]. %LAA values were stratified using three prespecified risk categories: ≤ 1, 1-5 and >5 [18]. %LAA and CAC were computed after convolutional neural network-based sharp to soft tissue kernel conversion [19].

### Statistical analysis

Study objectives were intra-subjects’ comparisons of CAC and %LAA values obtained with two different CT scanners in the inter-scanner cohort and with two different radiation dose CT protocols in the inter-dose cohort. Descriptive statistics were reported as numbers and percentages for categorical variables and as medians and interquartile ranges (IQR) for continuous variables. Agreement for CAC and %LAA categories was measured by the k-Cohen Index with Fleiss-Cohen weights (K_w_) and 95% Confidence Interval (CI). Correlations of continuous measurements were displayed by scatter plots and tested by Pearson’s Correlation Coefficients. Analyses were performed with Statistical Analysis System Software (SAS Studio 3.8, SAS Institute Inc., Cary, North Carolina, USA).

## Results

### Inter-scanner cohort

#### CAC

A total of 259 volunteers were tested with two different CT scanners; of them, 14 were excluded for software failure in CAC measurements and/or previous history of CVD (i.e., myocardial infarction, thrombosis, stroke, and angina) and/or of coronary stenting. The final cohort included 245 volunteers (Table 1). The median time between the two CTs was of 1.1 year (IQR 1-1.4). Median CAC score measured on the second-generation dual-source CT scanner was 59.3 (IQR 6.7-268.5); 144 volunteers (58.8%) scored 0-99, 56 (22.9%) 100-399 and 45 (18.4%) ≥ 400. Median CAC score measured on the third-generation dual-source CT scanner was 53.6 (IQR 3.3-273.4); 147 volunteers (60.0%) scored 0-99, 49 (20.0%) 100-399 and 49 (20.0%) ≥ 400. An overlap of CAC strata was observed in 218/245 (90%) volunteers with an almost perfect agreement: K_w_ values of 0.91 (95%CI 0.88-0.95). Second- and third-generation dual-source CT scanner CAC scores had a strongly positive correlation with Pearson’s Coefficient of 0.96 (p<0.001), as shown in Figure 1A.

**Table 1.** Comparison of CAC scoring in the inter-scanner cohort.

|  |  |  | *Third-generation dual source CT scanner* |  |  |  |
| --- | --- | --- | --- | --- | --- | --- |
|  |  |  | CAC score |  |  |  |
|  |  |  | 0-99 | 100-399 | ≥400 | Median (IQR) |
| Total volunteers |  | 245 | 147 (60.0%) | 49 (20.0%) | 49 (20.0%) | 53.6 (3.3-273.4) |
| *Second-generation dual source CT scanner*<br>CAC score | 0-99 | 144 (58.8%) | 137 (95.1%) | 7 (4.9%) | 0 | 6.5 (0.4-42.1) |
|  | 100-399 | 56 (22.9%) | 10 (17.9%) | 39 (69.6%) | 7 (12.5%) | 217.0 (114.7-292.2) |
|  | ≥400 | 45 (18.4%) | 0 | 3 (6.7%) | 42 (93.3%) | 742.1 (570.5-1243.6) |
|  | Median (IQR) | 59.3 (6.7-268.5) | 14.5 (2.2-51.9) | 207.1 (135.8-272.9) | 681.8 (488.1-1291.8) |  |

**Figure 1.**
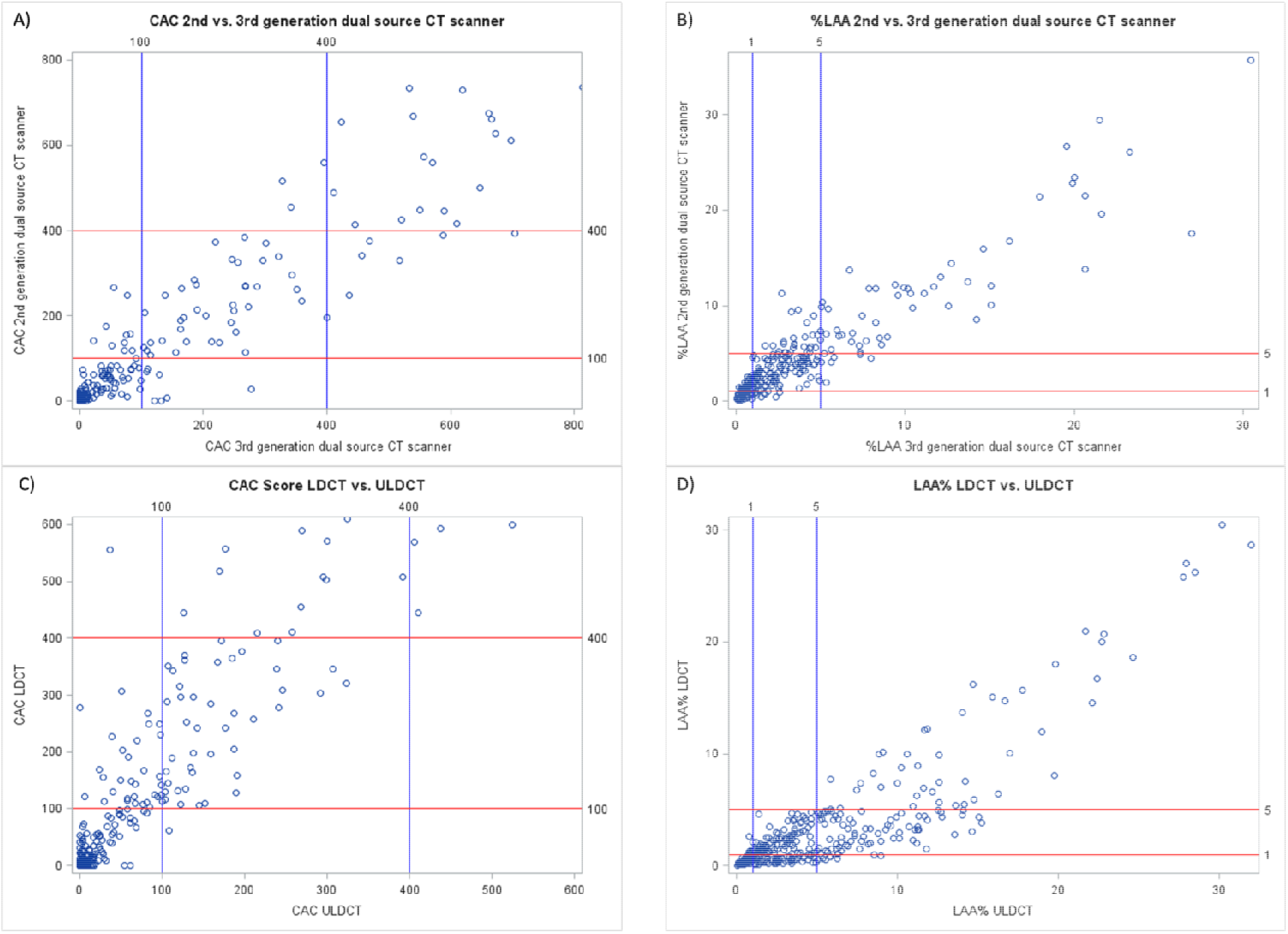

#### Emphysema

Three volunteers were excluded from the analysis due to software failure in %LAA measurements. The final cohort included 256 volunteers (Table 2). The median time between the two CTs was of 1.1 year (IQR 1-1.4).

**Table 2.** Comparison of %LAA measurements in the inter-scanner cohort.

|  |  |  | *Third-generation dual source CT scanner* |  |  |  |
| --- | --- | --- | --- | --- | --- | --- |
|  |  |  | %LAA |  |  |  |
|  |  |  | %LAA ≤ 1 | %LAA 1-5 | %LAA>5 | Median (IQR) |
| **Total volunteers** |  | **256** | **65 (25.4%)** | **134 (52.3%)** | **57 (22.3%)** | 2.5<br>(1.0-4.7) |
| *Second-generation dual source CT scanner*<br>**%LAA** | %LAA ≤ 1 | **43 (16.8%)** | 34 (79.1%) | 9 (20.9%) | 0 | 0.5<br>(0.2-0.9) |
|  | %LAA 1-5 | **136 (53.1%)** | 31 (22.8%) | 98 (72.1%) | 7 (5.2%) | 2.0<br>(1.1-3.6) |
|  | %LAA >5 | **77 (30.1%)** | 0 | 27 (35.1%) | 50 (64.9%) | 6.9<br>(4.4-12.1) |
|  | **Median (IQR)** | **3.4 (1.5-5.8)** | 1.0<br>(0.5-1.5) | 3.6<br>(2.4-4.7) | 10.0<br>(6.6-13.7) |  |

Median %LAA value measured on the second-generation dual-source CT scanner was 3.4 (IQR 1.5-5.8); 43 volunteers (16.8%) scored ≤ 1, 136 (53.1%) 1-5 and 77 (30.1%) >5.

Median %LAA value measured on the third-generation dual-source CT scanner was 2.5 (IQR 1.0-4.7), 65 volunteers (25.4%) scored ≤ 1, 134 (52.3%) 1-5 and 57 (22.3%) >5. An overlap of %LAA strata was observed in 182/256 (71%) volunteers, with a substantial agreement: K_w_ values of 0.70 (95%CI 0.63-0.76). Second-and third-generation dual-source CT scanner %LAA score had a strongly positive correlation with Pearson’s Coefficient of 0.92 (p<0.001), as shown in Figure 1B.

### Inter-dose cohort

#### CAC

Thirty-four volunteers were excluded due to software failure in CAC measurements and/or previous history of CVD (i.e., myocardial infarction, thrombosis, stroke, and angina) and/or of coronary stenting. The final cohort for CAC comparison analysis included 327 volunteers tested with LDCT and ULDCT protocols (Table 3).

**Table 3.** Comparison of CAC scoring in the inter-dose cohort.

|  |  |  | *ULDCT protocol*<br>CAC score |  |  |  |
| --- | --- | --- | --- | --- | --- | --- |
|  |  |  | 0-99 | 100-399 | ≥400 | Median (IQR) |
| Total volunteers |  | 327 | 219 (67.0%) | 61 (18.7%) | 47 (14.4%) | 27.3<br>(2.4-158.8) |
| *LDCT protocol*<br>CAC score | 0-99 | 187 (57.2%) | 186 (99.5%) | 1 (0.5%) | 0 | 3.7<br>(0.4-15.7) |
|  | 100-399 | 74 (22.6%) | 32 (43.2%) | 42 (56.8%) | 0 | 107.5<br>(77.0-158.8) |
|  | ≥400 | 66 (20.2%) | 1 (1.5%) | 18 (27.3%) | 47 (71.2%) | 589.6<br>(351.5-1265.2) |
|  | Median (IQR) | 53.8<br>(1.6-303.2) | 8.3<br>(0.2-54.8) | 315.0<br>(195.5-444.1) | 1089.4<br>(813.8-2241.2) |  |

Median value of CAC score on LDCT scan was 53.8 (IQR 1.6-303.2), 187 volunteers (57.2%) scored 0-99, 74 (22.6%) 100-399, and 66 (20.2%) ≥ 400.

Median value of CAC score on ULDCT scan was 27.3 (IQR 2.4-158.8), 219 volunteers (67.0%) scored 0-99, 61 (18.7%) 100-399, and 47 (14.4%) ≥ 400.

An overlap of CAC strata between LDCT and ULDCT scans was observed in 275/327 (84%) volunteers, with an almost perfect agreement: K_w_ of 0.86 (95%CI 0.82-0.90). LDCT and ULDCT CAC scores had a strongly positive correlation with Pearson’s Coefficient of 0.96 (p<0.001), as shown in Figure 1C.

Data on radiation exposure are reported in *Supplementary Table 1*.

#### Emphysema

Five screenees were excluded due to software failure in %LAA measurements. The final cohort for %LAA comparison analysis included 356 volunteers (Table 4).

**Table 4.**
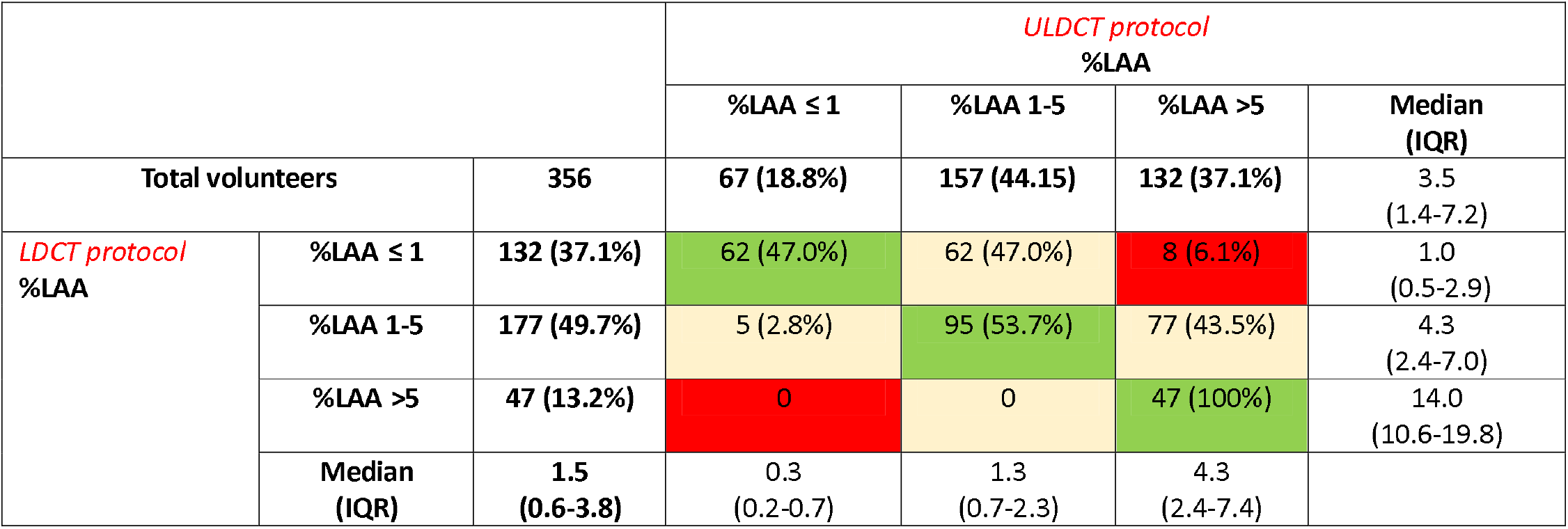
Comparison of %LAA measurements in the inter-dose cohort.

Median value of %LAA measured on LDCT was 1.5 (IQR 0.6-3.8), 132 volunteers (37.1%) scored ≤ 1, 177 (49.7%) 1-5, and 47 (13.2%) >5.

Median value of %LAA measured on ULDCT was 3.5 (IQR 1.4-7.2), 67 volunteers (18.8%) scored ≤ 1, 157 (44.1%) 1-5, and 132 (37.1%) >5.

An overlap of %LAA strata between LDCT and ULDCT scans was found in 204/356 (57%) volunteers, with a moderate agreement: K_w_ of 0.57 (95%CI 0.51-0.63). LDCT and ULDCT %LAA had a strongly positive correlation with Pearson’s Coefficient of 0.876 (p<0.001), as shown in Figure 1D.

Data on radiation exposure are reported in *Supplementary Table 1*.

## Discussion

We observed an overlap of CAC risk categories in 90% and 84% of screenees included in the inter-scanner and inter-dose cohort, respectively, with an almost perfect agreement, while the overlap for %LAA categories was overall lower, with a substantial agreement in the inter-scanner cohort and only moderate in the other cohort. These results are of particular interest given the burgeoning number of sites planning to start or already performing LCS activities. Reproducibility of measurements for CAC and emphysema intrinsically favor the applicability of LCS within national health systems, where different CT scanners and acquisition protocols are expected. Therefore, considering the possibility for a volunteer to be evaluated with different technical conditions, it is of paramount relevance to grant consistency of measurements between sites and scanners.

Discrepancies in the inter-scanner cohort can be partly due to the non-synchronous CT acquisition. Whether the slightly different CAC scores obtained for the two CT scanners were secondary to an actual change of CAC burden over time is difficult to say, being the time interval between the two acquisitions relatively short. Although the agreement was almost perfect (K_w_ of 0.86), an overlap of CAC risk categories was registered in a lower percentage of volunteers included in the inter-dose cohort as compared to the other cohort. A tendency towards a CAC score underestimation of ULDCT imaging was observed for 100-399 and ≥400 categories, whereas an opposite trend was noticed for the lowest risk category. Previous literature reported lower specificity and sensitivity of ULDCT imaging as compared to standard dose CT in this setting [20]. de Souza et al, however, did not assess the performance of an automated approach. Gorenstein et al, conversely, have recently tested a novel AI-based reconstruction denoising method for assessing both pulmonary nodules and CAC on ULDCT imaging in a LCS cohort of 123 subjects [21], demonstrating an accuracy of ULDCT imaging of 91.7% for CAC assessment.

As said for CAC, the differences observed in the inter-scanner cohort can be partly explained by the non-synchronous CT acquisition, being images acquired at potentially different inspiration levels. Although the time interval between the two LDCT scans acquisition was relatively short, making significant changes in emphysema extent rather unlikely, transient parenchyma abnormalities (e.g., small airways inflammation, atelectasis, etc.) might have affected emphysema quantification, and thus the comparison.

Albeit through different software and metrics, previous studies compared the detection of emphysema on ULDCT and LDCT imaging. Wang et al observed that iterative reconstruction (IR) algorithm improves the accuracy of ULDCT, reducing the overestimation of subtle emphysema, when compared to ULDCT scans reconstructed with analytic reconstruction algorithm, namely filtered back projection [22]. Kim et al tested two different ULDCT protocols (i.e., 100 kVp/20 mAs and 80 kVp/30 mAs) using IR in a non-LCS population, reporting that very low radiation doses should be avoided when emphysema is suspected [23]. The latter study, however, included a rather small number of subjects (25, of whom only 6 with emphysema), somehow limiting the significance of the observations. It is worth underlining that in the inter-dose cohort, the acquisition was performed during the same breath-hold, thus excluding potential variations caused by different inspiration levels, and images were reconstructed with an IR algorithm (ADMIRE), which was demonstrated to ensure higher images quality in terms of contrast-to-noise ratio (CNR) than that used by Kim et al (SAFIRE) [24].

Notably, some Authors have suggested that thresholds higher than -950HU (e.g., −940HU, −930HU) should be preferred for quantifying emphysema at LDCT imaging [25, 26]. As such, it can be speculated that a simple adjustment of software thresholds for emphysema quantification might overcome potential limitations due to different radiation dose CT protocols.

Our preliminary results suggest that a fully automated approach for CAC and emphysema quantification is feasible in the setting of LCS, whose continuous implementation at both national and international levels will likely pose the need for radiologists to analyze a significant number of heterogeneous images. Despite the differences that we observed among the two cohorts, mostly for emphysema extent, we believe that this automated approach can safely prompt further diagnostic investigations for CVD and COPD when either CAC or emphysema are depicted in LCS populations.

This study, however, has some limitations. First, as inherent to the nature of any retrospective study, our findings are subject to confounding factors. Second, the monocentric design with a single CT vendor reduces the generalizability of our results. Third, the non-synchronous CT acquisition in the inter-scanner cohort might have introduced a bias in the comparison of both CAC burden and emphysema extent. Forth, we only assessed the consistency of CAC score and %LAA categories between two different radiation dose CT protocols, but we did not test the diagnostic performance of ULDCT towards LDCT. Fifth, we did not assess potential effects of cholesterol-lowering therapies (e.g., statins) on the second evaluation in the inter-scanner cohort.

In conclusion, CAC and emphysema quantification performed by a fully automated AI software showed consistent results when applied on CT images acquired with different scanners and different radiation dose CT protocols in two LCS populations.

## Data Availability

All data produced in the present study are available upon reasonable request to the authors

## References

1. Heuvelmans, M.A., et al., Screening for Early Lung Cancer, Chronic Obstructive Pulmonary Disease, and Cardiovascular Disease (the Big-3) Using Low-dose Chest Computed Tomography: Current Evidence and Technical Considerations. J Thorac Imaging, 2019. 34(3): p. 160–169.

2. Yip, R., et al., Added benefits of early detection of other diseases on low-dose CT screening. Transl Lung Cancer Res, 2021. 10(2): p. 1141–1153.

3. Schreuder, A., et al., Combining pulmonary and cardiac computed tomography biomarkers for disease-specific risk modelling in lung cancer screening. Eur Respir J, 2021. 58(3).

4. de Koning, H.J., et al., Reduced Lung-Cancer Mortality with Volume CT Screening in a Randomized Trial. N Engl J Med, 2020. 382(6): p. 503–513.

5. Pastorino, U., et al., Ten-year results of the Multicentric Italian Lung Detection trial demonstrate the safety and efficacy of biennial lung cancer screening. Eur J Cancer, 2019. 118: p. 142–148.

6. Kauczor, H.U., et al., ESR/ERS statement paper on lung cancer screening. Eur Radiol, 2020. 30(6): p. 3277–3294.

7. Silvestri, G.A., et al., Outcomes From More Than 1 Million People Screened for Lung Cancer With Low-Dose CT Imaging. Chest, 2023.

8. Baldwin, D., et al., Developing a Pan-European Technical Standard for a Comprehensive High-quality Lung Cancer CT Screening Program. An ERS Technical Standard. Eur Respir J, 2023.

9. Blum, T.G., et al., European Respiratory Society guideline on various aspects of quality in lung cancer care. Eur Respir J, 2023. 61(2).

10. Balbi, M., et al., Automated Coronary Artery Calcium and Quantitative Emphysema in Lung Cancer Screening: Association With Mortality, Lung Cancer Incidence, and Airflow Obstruction. J Thorac Imaging, 2023. 38(4): p. W52–W63.

11. Alizadehsani, R., et al., Coronary artery disease detection using artificial intelligence techniques: A survey of trends, geographical differences and diagnostic features 1991-2020. Comput Biol Med, 2021. 128: p. 104095.

12. Fischer, A.M., et al., Artificial Intelligence-based Fully Automated Per Lobe Segmentation and Emphysema-quantification Based on Chest Computed Tomography Compared With Global Initiative for Chronic Obstructive Lung Disease Severity of Smokers. J Thorac Imaging, 2020. 35 Suppl 1: p. S28–S34.

13. Pastorino, U., et al., Baseline computed tomography screening and blood microRNA predict lung cancer risk and define adequate intervals in the BioMILD trial. Ann Oncol, 2022. 33(4): p. 395–405.

14. Milanese, G., et al., Ultra-low dose computed tomography protocols using spectral shaping for lung cancer screening: Comparison with low-dose for volumetric LungRADS classification. Eur J Radiol, 2023. 161: p. 110760.

15. van der Aalst, C.M., et al., Screening for cardiovascular disease risk using traditional risk factor assessment or coronary artery calcium scoring: the ROBINSCA trial. Eur Heart J Cardiovasc Imaging, 2020. 21(11): p. 1216–1224.

16. Bak, S.H., et al., Emphysema quantification using low-dose computed tomography with deep learning-based kernel conversion comparison. Eur Radiol, 2020. 30(12): p. 6779–6787.

17. Koo, H.J., et al., Prediction of Pulmonary Function in Patients with Chronic Obstructive Pulmonary Disease: Correlation with Quantitative CT Parameters. Korean J Radiol, 2019. 20(4): p. 683–692.

18. Labaki, W.W., et al., Quantitative Emphysema on Low-Dose CT Imaging of the Chest and Risk of Lung Cancer and Airflow Obstruction: An Analysis of the National Lung Screening Trial. Chest, 2021. 159(5): p. 1812–1820.

19. Lee, S.M., et al., CT Image Conversion among Different Reconstruction Kernels without a Sinogram by Using a Convolutional Neural Network. Korean J Radiol, 2019. 20(2): p. 295–303.

20. Silveira de Souza, V.V., et al., Performance of ultra-low-dose CT for the evaluation of coronary calcification: a direct comparison with coronary calcium score. Clin Radiol, 2017. 72(9): p. 745–750.

21. Gorenstein, L., et al., A Novel Artificial Intelligence Based Denoising Method for Ultra-Low Dose CT Used for Lung Cancer Screening. Acad Radiol, 2023. 30(11): p. 2588–2597.

22. Wang, R., et al., Ultralow-radiation-dose chest CT: accuracy for lung densitometry and emphysema detection. AJR Am J Roentgenol, 2015. 204(4): p. 743–9.

23. Kim, Y., et al., Ultra-Low-Dose CT of the Thorax Using Iterative Reconstruction: Evaluation of Image Quality and Radiation Dose Reduction. AJR Am J Roentgenol, 2015. 204(6): p. 1197–202.

24. Shin, J.B., et al., Comparative performance analysis for abdominal phantom ROI detectability according to CT reconstruction algorithm: ADMIRE. J Appl Clin Med Phys, 2020. 21(1): p. 136–143.

25. den Harder, A.M., et al., Emphysema quantification using chest CT: influence of radiation dose reduction and reconstruction technique. Eur Radiol Exp, 2018. 2(1): p. 30.

26. Cao, X., et al., Optimal threshold in low-dose CT quantification of emphysema. Eur J Radiol, 2020. 129: p. 109094.

