## Supplementary for "Consistency of automated coronary calcium score and extent of emphysema with different CT scanners and radiation dose protocols in lung cancer screening"

### Supplementary materials

#### Scanning acquisition and reconstruction parameters

##### *Somatom Flash*

The whole chest volume was scanned on cranio-caudal direction, during one deep inspiratory breath-hold with the following scanning parameters: tube voltage, 120 kVp; tube current, 30 mAs; collimation, 0.625 mm; pitch, 1.2; rotation time, 0.5 seconds. Images were reconstructed with the following parameters: thickness, 1 mm; increment, 0.7 mm; medium-sharp kernel (B50f).

##### *Somatom Force*

LDCT scans were acquired on cranio-caudal direction, during one deep inspiratory breath-hold with the following scanning parameters: tube voltage, 120 kVp; tube current, 25 mAs; while ULDCT scans on caudo-cranial direction using a spectral shaping by tin filter and one of the following protocols [1]:

1. ULDCT_1_: fully automated approach for modulation of tube potential (CARE kV, Sn100kVp – Sn150 kVp) and automated exposure control (AEC for current at reference 100mAs);

2. ULDCT_2_: fixed tube-voltage and tube current according to patient size (Sn100kVp; 140mAs for screenees whose field of view was 300 mm, 210mAs for screenees whose FOV was 350/400 mm);

3. ULDCT_3_: hybrid approach (fixed tube-voltage at Sn100kVp and AEC for current at reference 100mAs);

4. ULDCT_4_: hybrid approach (fixed tube-voltage at Sn150kVp and AEC for current at reference 20mAs).

LDCT and ULDCT were reconstructed with the following parameters: slice thickness of 1 mm, increment of 0.7 mm. A medium-sharp kernel (Br49) and advanced modelled iterative reconstruction (IR, ADMIRE strength level 3) were applied to LDCT scans, whereas two medium-sharp kernels and IR strength levels to ULDCT scans: Br49 (ADMIRE 3) and Qr49 (ADMIRE 4).

#### Radiation dose

Dose-Length Product (DLP) and Volume CT dose index (CTDIvol) were recorded for LDCT and ULDCT; effective doses (ED) were estimated using the conversion coefficient of 0.014 [2].

1.	Milanese, G., et al., *Ultra-low dose computed tomography protocols using spectral shaping for lung cancer screening: Comparison with low-dose for volumetric LungRADS classification.* Eur J Radiol, 2023. **161**: p. 110760.

2.	Deak, P.D., Y. Smal, and W.A. Kalender, *Multisection CT protocols: sex- and age-specific conversion factors used to determine effective dose from dose-length product.* Radiology, 2010. **257**(1): p. 158-66.
